# Regional healthy brain activity, glioma occurrence and symptomatology

**DOI:** 10.1101/2022.01.14.22269290

**Authors:** Tianne Numan, Lucas C. Breedt, Bernardo de A.P.C. Maciel, Shanna D. Kulik, Jolanda Derks, Menno M. Schoonheim, Martin Klein, Philip C. de Witt Hamer, Julie J. Miller, Elizabeth R. Gerstner, Steven M. Stufflebeam, Arjan Hillebrand, Cornelis J. Stam, Jeroen J.G. Geurts, Jaap C. Reijneveld, Linda Douw

## Abstract

It is unclear why exactly gliomas show preferential occurrence in certain brain areas. Increased spiking activity around gliomas leads to faster tumor growth in animal models, while higher non-invasively measured brain activity is related to shorter survival in patients. However, it is unknown how regional intrinsic brain activity, as measured in healthy controls, relates to glioma occurrence. We first investigated whether gliomas occur more frequently in regions with intrinsically higher brain activity. Secondly, we explored whether intrinsic cortical activity at individual patients’ tumor locations relates to tumor and patient characteristics.

Across three cross-sectional cohorts, 306 patients with different tumor subtypes were included. Tumor masks were created for each patient. Intrinsic regional brain activity was assessed through resting-state magnetoencephalography (MEG) acquired in healthy controls, which was then source-localized to 210 cortical brain regions according to the Brainnetome atlas. Brain activity was operationalized as (1) broadband power and (2) offset of the aperiodic component of the power spectrum, which both reflect neuronal spiking of the underlying neuronal population. We additionally assessed (3) the slope of the aperiodic component of the power spectrum, which is thought to reflect the neuronal excitation/inhibition (E/I) balance. First, correlation coefficients were calculated between group-level regional glioma occurrence, as obtained by concatenating tumor masks across patients, and group-averaged regional intrinsic brain activity. Second, intrinsic brain activity at specific tumor locations was calculated by overlaying patients’ individual tumor masks with regional intrinsic brain activity of the controls and was associated with tumor and patient characteristics.

As hypothesized, more malignant glioma preferentially occurred in brain regions characterized by higher intrinsic brain activity in controls as reflected by higher offset. Secondly, intrinsic brain activity at patients’ individual tumor locations differed according to glioma subtype and performance status: the most malignant IDH-wildtype glioblastoma patients had the lowest E/I balance at their individual tumor locations, while lower E/I balance weakly related to poorer Karnofsky performance status, independent of glioma subtype.

In conclusion, malignant gliomas more frequently occur in cortical brain regions with intrinsically higher activity levels, suggesting that more active regions are more vulnerable to glioma development. Moreover, indices of healthy, intrinsic E/I balance at patients’ individual tumor locations may capture both tumor biology and patients’ performance status. These findings contribute to our understanding of the complex and bidirectional relationship between normal brain functioning and glioma growth, which is at the core of the relatively new field of ‘cancer neuroscience’.

## Introduction

Glioma, the most frequently occurring primary brain tumor, unequivocally impacts functioning of the rest of the brain. However, seemingly normal variations in brain functioning also affect how gliomas behave, as preclinical work in animal models of glioma as well as *in vitro* studies have shown. Higher activity of neurons surrounding high-grade gliomas causes an acceleration of glioma growth through glutamate-dependent “neurogliomal synapses”.^1–6^ Development of these synapses is promoted through increased neuroligin-3 (NLGN3) expression, which is also caused by higher neuronal activity.^2,3,7^ In three glioblastoma patients undergoing electrocorticography during tumor resection, brain activity as measured by gamma band power was significantly higher around the tumor than in more distant regions of the brain.^2^ Similar results have also been observed when using broadband power measured non-invasively with magnetoencephalography (MEG) and electroencephalography (EEG) as a proxy of neuronal activity.^8^ Furthermore, there are indications that healthy and pathological activity in the (peri)tumoral region are interdigitated during cognitive performance in these patients.^9^ This complex bidirectional interaction between the brain and glioma that informs both tumor and patient behavior has sparked the relatively new field of ‘cancer neuroscience’, aiming to elucidate how this crosstalk may inform understanding and treatment of glioma.^10,11^

A potential implication of this line of research pertains to the relevance of ‘intrinsic’ brain activity, i.e. the assumed preexisting level of brain activity as measured in healthy controls, for glioma occurrence. The distribution of glioma across the neocortex is not random: there is a preponderance of occurrence in the gray matter of the frontal and temporal lobes, with occipital tumors being rare.^12,13^ Intrinsic regional brain functioning, as measured in healthy subjects, may therefore hold clues as to why gliomas preferentially occur in those brain areas. A recent study used resting-state functional MRI (fMRI) network indicators of intrinsic regional hubness (i.e. the relative importance of regions in the brain network) to show that glioma most often occurs in those areas more functionally connected to other brain regions.^14^ Moreover, gliomas seem to preferentially occur in brain regions normally characterized by greater MEG network clustering.^15^ Interestingly, work in other neurological diseases has shown that lesions occurring in such highly connected, putatively also highly active,^16^ brain regions have a bigger impact on global brain network functioning as well as neurological symptomatology than lesions in less-connected areas of the brain.^17–19^ However, it is unknown how regional intrinsic brain activity relates to glioma occurrence, and how activity at individual tumor locations may concurrently pertain to pathophysiological characteristics, such as glioma subtype and patients’ performance status. The latter may be of particular interest, since recent work suggests that the interplay between peritumoral neurons and glioma cells relates to both cognitive functioning and survival.^20^

Combining these lines of research, we here set out to first test whether intrinsic, healthy regional variance in brain activity could further increase our understanding of preferential glioma occurrence. We measured brain activity non-invasively in a cohort of healthy controls with MEG and calculated three proxies of neuronal activity per brain region. Based on the relationship between spiking activity and tumor growth, we hypothesized gliomas to preferentially occur in intrinsically more active brain areas, particularly so for more malignant gliomas for which the relationship between neuronal activity and proliferation has been amply shown in preclinical studies. Second, we aimed to use intrinsic brain activity at each individual patient’s tumor location as a correlate of tumor and patient characteristics, in order to investigate whether it would reflect the tumor biology, symptomatology and prognosis of these patients. If so, such findings could lead to the discovery of treatment targets for this lethal disease.

## Materials and methods

### Patient cohorts

Three newly diagnosed diffuse glioma patient cohorts were used in this study. The Amsterdam (AMS) cohort was selected retrospectively from a set of patients who visited the Brain Tumor Center Amsterdam of the Amsterdam University Medical Centers (Amsterdam, The Netherlands) between 2004-2013 with a suspected diffuse glioma. This cohort participated in previous work and consisted of patients undergoing clinical task-based fMRI for motor and/or language localization before tumor resection.^21^ Further inclusion criteria for the study were age of 18 years or older and histopathologically confirmed diffuse glioma according to the World Health Organization (WHO) 2007 classification.^22^ Inclusion of these patients and use of their data was approved by the Medical Ethical Committee of the VU University Medical Center.

The second cohort was also selected retrospectively, from the patient population visiting Massachusetts General Hospital (Boston, MA, United States of America; MGH cohort) between 2006-2013 with suspected diffuse glioma. These patients were included in previous work and consisted of patients undergoing anatomical and diffusion MRI.^23^ Further inclusion criteria were identical to the AMS cohort. Inclusion of these patients and use of their data was approved by the Institutional Review Board of the Dana Farber Cancer Institute.

Finally, we used the publicly available glioblastoma dataset from The Cancer Genome Atlas combined with The Cancer Imaging Archive (TCGA-TCIA) as a source of tumor masks drawn by a different expert panel; these tumor masks were also validated through the Brain Tumor Segmentation (BraTS) challenge.^24^

### Patient and tumor-related characteristics

Medical chart review was used to extract additional relevant patient characteristics for the AMS and MGH cohorts at the preoperative timepoint, which was concurrent to the MRI scan on which tumor masks were drawn in. These characteristics included age, sex, Karnofsky Performance Status (KPS^25^), presence or absence of epilepsy, and disease course. Medical chart review also revealed clinically determined tumor grade and molecular subtype (i.e. isocitrate dehydrogenase (IDH) and 1p/19q status), according to the 2016 WHO classification.^26^ Three main molecular subgroups were defined: (1) IDH-mutant, 1p/19q-codeleted (grade II and III) gliomas, (2) IDH-mutant (grade II and III) gliomas without 1p/19q codeletion, and (3) IDH-wildtype (grade IV) glioblastomas. IDH-mutant glioblastomas and IDH-wildtype grade II and III gliomas are rare and were thus excluded from subgroup analyses.

For each individual patient, progression-free survival (PFS) was defined as the time in weeks between date of the preoperative MRI and date of the moment of progression, clinically defined by the treating physician and/or the multidisciplinary tumor board including neurologists, neurosurgeons, neuroradiologists, radiation oncologists and medical oncologists. Overall survival (OS) was defined as the time between the preoperative MRI and death in weeks. Patients without progression or death were censored at the time of their last contact with the treating neuro-oncologist.

For the TCGA dataset, only tumor masks, sex and (in a subset of cases) age were available.

### Glioma occurrence maps

All information on the TCGA dataset and tumor masks can be found elsewhere.^24^ In the AMS and MGH cohorts, preoperative anatomical MRI was obtained. All tumor masks were manually drawn using the preoperative (3D)T1 with and without contrast injection and fluid-attenuated inversion recovery (FLAIR) images, as is the current standard in clinical neuro-oncology.^27^ Tumor borders were largely determined using the contrast-enhancing tumor rims, unless FLAIR images showed more extensive tumor infiltration beyond contrast enhancement, in which cases the tumor mask was extended to also incorporate these FLAIR hyperintense regions. In non-enhancing glioma, a combination of T1 hypointensities (tumor core) and FLAIR hyperintensities (tumor infiltration) was used to draw in tumor masks. Tumor masks were then linearly transformed to MNI standard space based on brain extracted scans using FSL’s FLIRT with nearest neighbor interpolation and 12 degrees of freedom (FSL 5.0.10).^28^

Glioma masks were then merged into a tumor occurrence heat map per cohort or per cohort-specific subgroup, representing the relative occurrence of tumor per voxel. This voxel-based map was then averaged across the 210 cortical regions of the Brainnetome atlas^29^ to create a regional map of tumor occurrence, as schematically depicted in Figure 1.

**Figure 1.**
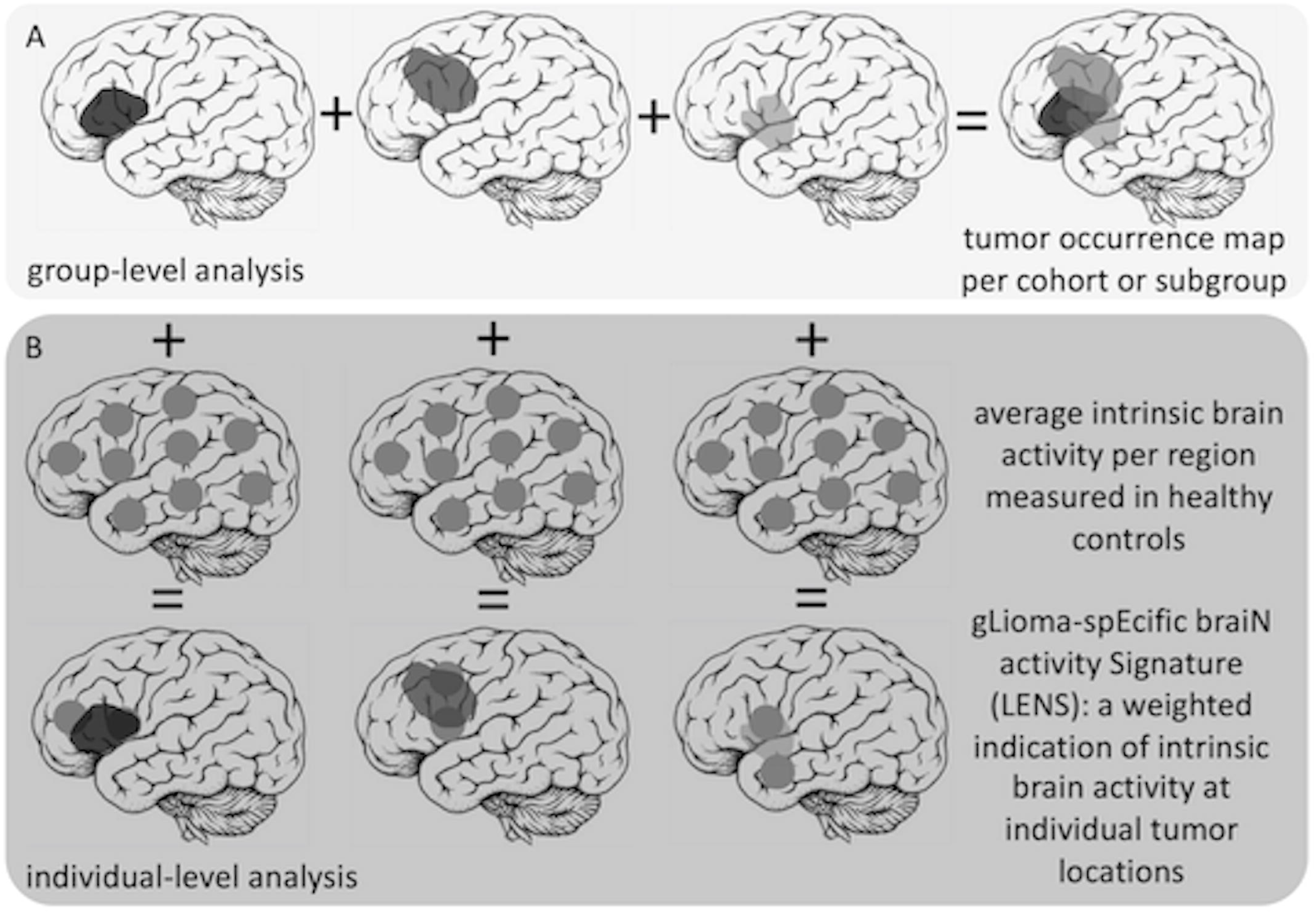
Schematic overview of patient-related analyses Legend. In (A), the construction of the tumor occurrence map per cohort or subgroup by summation of all tumor masks in template space is depicted. In (B), individual tumor masks are overlaid with regional brain activity values of healthy controls. Brain activity values are then weighted according to percent overlap with the tumor, and the ultimate average gLioma-spEcific braiN activity Signature (LENS) is normalized for the total number of regions overlapping with the tumor.

### Intrinsic regional brain activity

Healthy subjects were selected from a cohort previously collected as a control group in non-glioma studies,^30–32^ consisting of 65 healthy controls who underwent, amongst others, structural MRI and MEG. From this cohort, controls were selected by initially matching them to the AMS patient cohort at the group-level with respect to age and sex. Inclusion of these subjects was approved by the VU University Medical Center ethical review board and all subjects gave written informed consent before participation.

Healthy controls underwent 5-minute eyes-closed MEG in supine position using a whole-head system (Elekta Neuromag Oy, Helsinki, Finland) with 306 channels inside a magnetically shielded room (Vacuumschmelze GmbH, Hanau, Germany). Data were recorded with a sampling frequency of 1250Hz, filtered online with a 410Hz anti-aliasing filter and a 0.1Hz high-pass filter. The head position relative to the sensors was recorded continuously with head-localization coils. A 3D digitizer (Fastrak, Polhemus, Colchester, VT, USA) digitized the head-localization coil positions and scalp outline (roughly 500 points) to allow surface matching of the scalp surface points with anatomical MRI.

These data were processed as previously described.^33–35^ In short, data were visually inspected and at maximum 12 malfunctioning channels were excluded. Offline spatial filtering of the raw data removed artifacts using the temporal extension of Signal Space Separation (tSSS) using MaxFilter software (Elekta Neuromag Oy; version 2.1).^36^ The reconstruction of neuronal sources was performed with an atlas-based beamforming approach^37^ that uses weight normalization, after which time series (virtual electrodes) for each centroid of each atlas region were reconstructed in the broadband frequency range (0.5-48Hz).^38^ Ten artifact-free epochs of 13.3 seconds each were used for further analysis.

Three measures of brain activity were then calculated using these epochs: (1) broadband power of the power spectrum, (2) offset, and (3) slope of the aperiodic component of the power spectrum, as described before.^8^ Broadband power reflects the spectral density or area under the curve of the total spectral signal measured with MEG, and reflects the spiking rates of the underlying neuronal populations.^39,40^ The offset is related to broadband power, but filters out frequency-specific variations in signals to establish a potentially more specific measure of neuronal spiking.^41^ Finally, the slope takes into account the distribution of signal across the entire frequency spectrum, with a steeper slope (i.e. more low-frequency signal) reflecting a lower E/I balance,^41^ thus indicating relatively more inhibition occurring in the underlying neuronal populations. This measure, although not a direct reflection of neuronal activity, was taken into account as well, since *in vitro* experiments on glioma tissue have shown that blockage of excitatory AMPA receptors through the anti-seizure medication perampanel inhibits tumor growth in the context of increased neuronal spiking, supporting the idea that the balance between excitation and inhibition may be of relevance in glioma, also for future treatment strategies.^1^ These three measures are interrelated: broadband power and offset are most straightforwardly related to each other, while the relationship between slope and the other two measures is less evident. Previous studies in healthy controls have only investigated the relationship between band-specific power and slope, such that greater alpha power goes hand in hand with relatively less steep slope values (higher excitation over inhibition) when also determined over a low frequency range.^41,42^ It remains unknown how broadband slope associates with power and offset.

First, for each epoch and each brain region, the power spectrum was obtained, using Welch’s method with four non-overlapping Hamming windows, and averaged across the ten epochs. Broadband power was then calculated as the area under the power spectral density between 0.5-48 Hz for each brain region. The offset and slope of each power spectral density was obtained by using an automated parameterizing algorithm (FOOOF version 0.1.3, an open source package (https://github.com/fooof-tools/fooof/ implemented in Python v3.7.0)).^41^ The power spectrum, *P,* was modelled using three parameters:

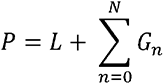

where *L* is the aperiodic “background” signal, with *N* total peaks extracted from the power spectrum, and Gaussians (*G_n_*) fitted to each peak. The peaks were iteratively fitted by Gaussians:

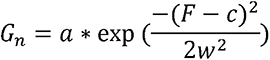

With *a* amplitude, *c* center frequency, *w* the bandwidth of the Gaussian *G*, and *F*, the input frequencies. The aperiodic signal L was modelled by:

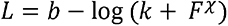

Where *b* is the broadband offset, χ is the slope, and *k* is the “knee” parameter which was set to 0. The FOOOF model was fitted for the frequency range 0.5-48 Hz.

Broadband power, offset and slope were then averaged across all healthy subjects to obtain an average distribution of values across the 210 cortical regions. We then excluded the medial brain regions, as they are further away from the MEG sensors and thus have low signal-to-noise (SNR) ratios, potentially confounding any results (see Supplementary Figure 1).^43,44^ Additional analyses taking all regions into account can be found in the Supplementary Materials.

### Intrinsic brain activity signatures at individual tumor locations

In order to assess the clinical relevance of healthy, intrinsic brain activity at patients’ individual glioma locations, a gLioma-spEcific braiN activity Signature (LENS) was calculated (Figure 1). Each tumor mask was overlaid with the group-averaged intrinsic brain activity value (broadband power, offset or slope) per region. We multiplied the percentage of each brain region invaded by a patient’s glioma with the healthy brain activity value for that same region, then summed these values over all tumor-invaded brain regions, and finally divided the sum by the number of regions invaded by tumor. This yielded three LENS values per patient that were corrected for tumor size: LENS-bbp (broadband power), LENS-offset, and LENS-slope.

### Statistical analysis

Statistical analyses were performed with Matlab (version R2020.b, Mathworks, Natick, MA, USA) and IBM SPSS Statistics (version 22, IBM Corp., Armonk, NY, USA). Data are presented as mean (standard deviation (SD)) when normally distributed, or median (1^st^ quartile (Q1) – 3^rd^ quartile (Q3)) when deviating from a normal distribution. Group differences in age and sex were explored using Student’s t-tests and chi-square tests.

Regional glioma occurrence was correlated with group-averaged regional healthy brain activity of all included cortical regions using Spearman’s correlation coefficients, within all cohorts and subgroups. For these analyses, we considered a p-value below 0.0019 as significant, using Bonferroni correction to account for the 27 tests performed.

Since correlations between brain maps may in part be due to the higher chance that neighboring regions display comparable variations, we additionally performed the spin test using publicly available code to do so (https://www.github.com/spin-test).45 This procedure uses a spatial permutation framework to generate null models of correlation by applying random rotations to spherical projections of both regional glioma occurrence and regional brain activity mapped onto the cortical surface. The non-cortical regions (medial wall, subcortical areas) were excluded. For any significant results, we additionally checked their robustness using these null models to create a distribution of correlation coefficients.

Subgroup differences in LENS values were tested using Kruskal-Wallis signed rank tests (in the AMS cohort with three subgroups) followed by post-hoc Dunn pairwise tests in case of significant results, and with Mann-Whitney U tests (in the MGH cohort with two subgroups, and for post-hoc testing within the AMS cohort). Associations between LENS values and tumor volume were tested using Spearman’s correlation coefficients. Differences in LENS values according to presence of epilepsy and low versus high KPS, were tested using Mann-Whitney U tests. Finally, univariate Cox proportional hazards analyses of the relationships between LENS values and OS and PFS were performed. These analyses were not corrected for multiple comparisons, a p-value below 0.05 was considered significant.

### Data availability

The derivative data and code used in this manuscript are available for download at https://github.com/multinetlab-amsterdam/projects/.

## Results

### Subject characteristics

Characteristics of the main patient cohorts and subgroups can be found in Table 1, and tumor occurrence maps per cohort displayed the expected preference of glioma to occur in the frontoparietal regions (Supplementary Figure 2). The AMS cohort consisted mainly of IDH-mutant gliomas, while the MGH cohort consisted mostly of IDH-wildtype glioblastomas, and the TCGA cohort consisted only of IDH-wildtype glioblastomas. Subgroup characteristics reflected the expected differences between these cohorts: the AMS cohort was younger than both other cohorts, with AMS patients generally having longer survival than the MGH cohort.

**Table 1.** Patient characteristics

| Subgroup | AMS cohort |  |  |  |
| --- | --- | --- | --- | --- |
|  | all patients (n=83) | subgroup 1 (n=21) | subgroup 2 (n=30) | subgroup 3 (n=15) |
| Mean age in years (SD) | 41 (12) | 43 (9) | 35 (10) | 54 (13) |
| Number of males \| females | 55 \| 28 | 14 \| 7 | 16 \| 14 | 13 \| 2 |
| Handedness right \| left \| unknown | 42 \| 8 \| 33 | 10 \| 2 \| 9 | 15 \| 4 \| 11 | 9 \| 0 \| 6 |
| Glioma localization left \| right | 46 \| 37 | 12 \| 9 | 20 \| 10 | 9 \| 6 |
| Glioma grade II \| III \| IV | 46 \| 22 \| 15 | 9 \| 12 \| 0 | 20 \| 10 \| 0 | 0 \| 0 \| 15 |
| Number of patients with IDH-1p/19q status unknown (%) | 17 (21%) | NA | NA | NA |
| Median tumor volume in cm<sup>3</sup> in MNI space (Q1-Q3) | 89 (43-158) | 79 (47 - 169) | 88 (45-140) | 117 (52-187) |
| Median PFS in weeks (Q1-Q3) | 106 (42-223) | 235 (92-286) | 79 (45-224) | 42 (18-66) |
| Median OS in weeks (Q1-Q3) | 167 (71-321) | 277 (180-452) | 268 (213-323) | 70 (49-114) |
| Number of patients with KPS < 90 \| 90-100 \| unknown | 23 \| 38 \| 22 | 7 \| 10 \| 4 | 5 \| 18 \| 7 | 5 \| 3 \| 7 |
| Number of patients with epilepsy \| without epilepsy \| unknown | 60 \| 11 \| 12 | 14 \| 3 \| 4 | 24 \| 3 \| 3 | 10 \| 3 \| 2 |
| Subgroup | BOS cohort |  |  |  |
|  | all patients (n=121) | NA | subgroup 2 (n=18) | subgroup 3 (n=91) |
| Mean age in years (SD) | 53 (14) |  | 41 (14) | 57 (12) |
| Number of males \| females | 67 \| 54 |  | 7 \| 11 | 54 \| 37 |
| Glioma localization left \| right \| bilateral | 58 \| 60 \| 1 |  | 9 \| 8 | 46 \| 44 \| 1 |
| Glioma grade II \| III \| IV | 19 \| 8 \| 94 |  | 11 \| 7 | 0 \| 0 \| 91 |
| Number of patients with IDH-1p/19q status unknown (%) | 2 (2%) |  | NA | NA |
| Median tumor volume in cm<sup>3</sup> in MNI space (Q1-Q3) | 34 (18-64) |  | 46 (18-88) | 32 (19-56) |
| Median PFS in weeks (Q1-Q3; n with progression) | 46 (33-77; 73) |  | 85 (30-149; 6) | 44 (33-65; 63) |
| Median OS in weeks (Q1-Q3; n died) | 79 (48-119; 58) |  | NA | 79 (48-120; 56) |
| Number of patients with KPS < 90 \| 90-100 \| unknown | 30 \| 77 \| 14 |  | 4 \| 14 | 25 \| 52 \| 14 |
| Number of patients with epilepsy \| without epilepsy \| unknown | 69 \| 22 \| 30 |  | 12 \| 5 \| 1 | 57 \| 17 \| 17 |

| Subgroup | TCGA cohort |  |  | subgroup 3<br>(n=102) |
| --- | --- | --- | --- | --- |
|  | NA | NA | NA |  |
| Mean age in years (SD), known in n=63 |  |  |  | 57 (15) |
| Number of males \| females |  |  |  | 66 \| 36 |

Legend. AMS = Amsterdam, BOS = Boston, TCGA = The Cancer Genome Atlas, SD = standard deviation, PFS = progression-free survival, OS = overall survival, KPS = Karnofsky Performance Stauts, MNI = Montreal Neurological Institute.

In total, 45 of 65 available healthy controls were matched primarily to the AMS patient cohort, since we had most extensive clinical information on this cohort. The matched healthy cohort consisted of 25 males (20 females), with a mean age of 43 years (SD = 9). As could be expected based on the literature, regional broadband power and offset values were most strongly correlated (Spearman’s rho = 0.734, *P* < 0.001), followed by a correlation between offset and slope values (rho = -0.312, *P* < 0.001) and finally between broadband power and slope (rho = - 0.299, *P* < 0.001).

### Associations between regional glioma occurrence and intrinsic brain activity

We first set out to test the hypothesis that glioma preferentially occurs in brain regions that are characterized by intrinsically greater brain activity. Indeed, higher regional healthy brain activity values in terms of offset were significantly and robustly related to higher regional glioma occurrence across cohorts with respect to IDH-mutant, non-codeleted tumors and IDH-wildtype glioma (rho > 0.309, *P* < 0.0019, Table 2, Figure 2). For IDH-wildtype tumors, there was also a robust association with intrinsic slope, in addition to significant associations with offset and broadband power (rho < -0.387, *P* < 0.0019). The smallest subgroup of IDH-mutant, 1p/19q-codeleted tumor occurrence maps did not show any significant associations with intrinsic brain activity measures.

**Figure 2.**
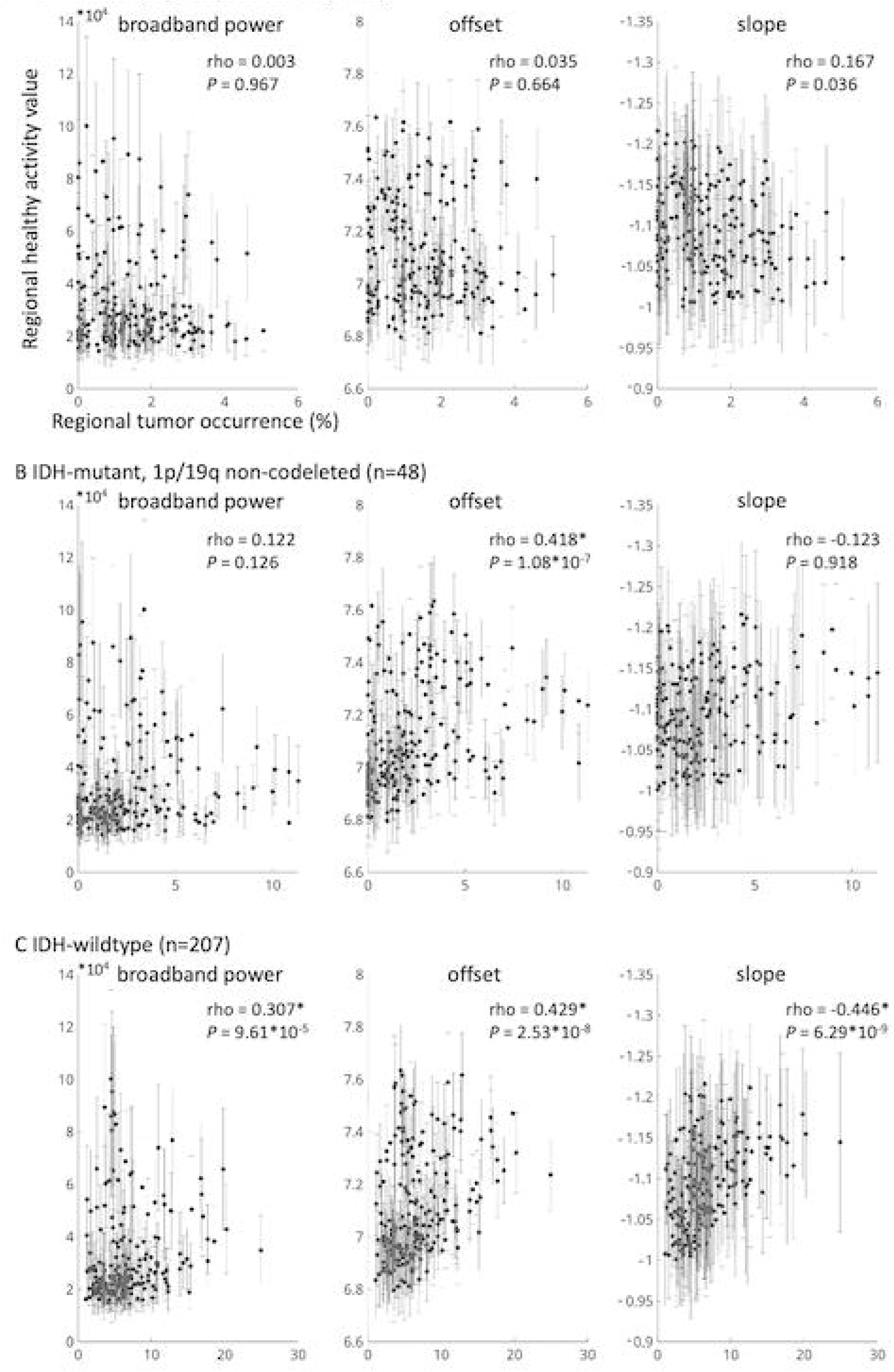
Correlations between tumor occurrence and intrinsic regional brain activity per tumor subtype Legend. (A) shows the correlation between regional tumor occurrence of all IDH-mutant, 1p/19q-codeleted gliomas on the x-axis and average regional intrinsic brain activity of the healthy controls on the y-axis (error bars indicate standard deviation per region), for broadband power (left), offset (middle), and slope (right). Axis titles are identical for all plots. (B) and (C) show the same correlations for all IDH-mutant, 1p/19q non-codeleted gliomas and IDH-wildtype gliomas, respectively. * = statistically significant after correction for the total number of tests performed.

**Table 2.** Associations between regional glioma occurrence and regional intrinsic brain activity

| Subgroup | Cohort | N | Broadband power | Offset | Slope |
| --- | --- | --- | --- | --- | --- |
| IDH-mutant, 1p/19q-codeleted glioma | AMS | 21 | 0.003 | 0.035 | 0.167 |
| IDH-mutant, non-codeleted glioma | AMS | 30 | 0.111 | 0.396** | -0.069 |
|  | BOS | 18 | 0.062 | 0.305* | 0.089 |
|  | *combined* | 48 | 0.122 | 0.407** | -0.008 |
| IDH-wildtype glioma | AMS | 14 | 0.177 | 0.418* | -0.123 |
|  | BOS | 91 | 0.278** | 0.315** | -0.437** |
|  | TCGA | 102 | 0.270** | 0.309** | -0.387** |
|  | *combined* | 207 | 0.307** | 0.429** | -0.446** |

\* $P < 0.0019$ , significant after correction for multiple comparisons across 27 tests, \*\* $P < 0.0019$ after multiple comparisons correction and significant when using the spin test to create null models. AMS = Amsterdam, BOS = Boston, TCGA = The Cancer Genome Atlas. Values indicate Spearman correlation coefficients.

Several additional analyses were performed to check the robustness of these results (see Supplementary Materials for details). All significant correlations remained significant after creating null models of spatial correlations using the spin test (Table 2), except for the correlation between offset and tumor occurrence in the AMS (*P*_spin_ = 0.051) and MGH (*P*_spin_ = 0.137) cohorts (see Supplementary Table 1 for further details). Moreover, since the included healthy controls were significantly younger than patients in the MGH and TCGA cohorts, additional analyses were performed to assure that any findings in the MGH/TCGA cohorts were not due to this age difference. Results did not change when considering the medial regions as well (Supplementary Table 2). Moreover, since we used the average of the healthy controls per region as an indication of intrinsic brain activity, we explored whether regional differences were larger than individual variations in our three measures. This largely proved to be the case (Supplementary Figure 1), further supporting the robustness of these findings.

### LENS differences between molecular and performance status subgroups

We then investigated whether LENS, the gLioma-spEcific braiN activity Signature (i.e. the weighted value characterizing intrinsic healthy brain activity at individual patients’ glioma locations) differed per subgroup. In the AMS cohort, LENS-slope differed between the three subgroups (Kruskal-Wallis Z = 8.77, *P* = 0.012, Figure 3), but LENS-bbp (Z = 0.25, *P* = 0.291) and LENS-offset (Z = 4.53, *P* = 0.104) did not. Post-hoc tests revealed that the IDH-mutant, 1p/19q-codeleted subgroup had significantly less steep LENS-slopes than the IDH-mutant, non-codeleted patients (Dunn statistic = -11.6, *P* = 0.034) and IDH-wildtype patients (Dunn statistic = -18.5, *P* = 0.004). In the MGH cohort, these results were partly replicated: LENS-slope was less steep in the IDH-mutant, non-codeleted glioma patients as compared to the IDH-wildtype glioblastoma patients (Figure 3; Mann-Whitney U = 547, *P* = 0.026), while LENS-bbp (U = 662, *P* = 0.200) and LENS-offset (U = 760, *P* = 0.630) did not differ.

**Figure 3.**
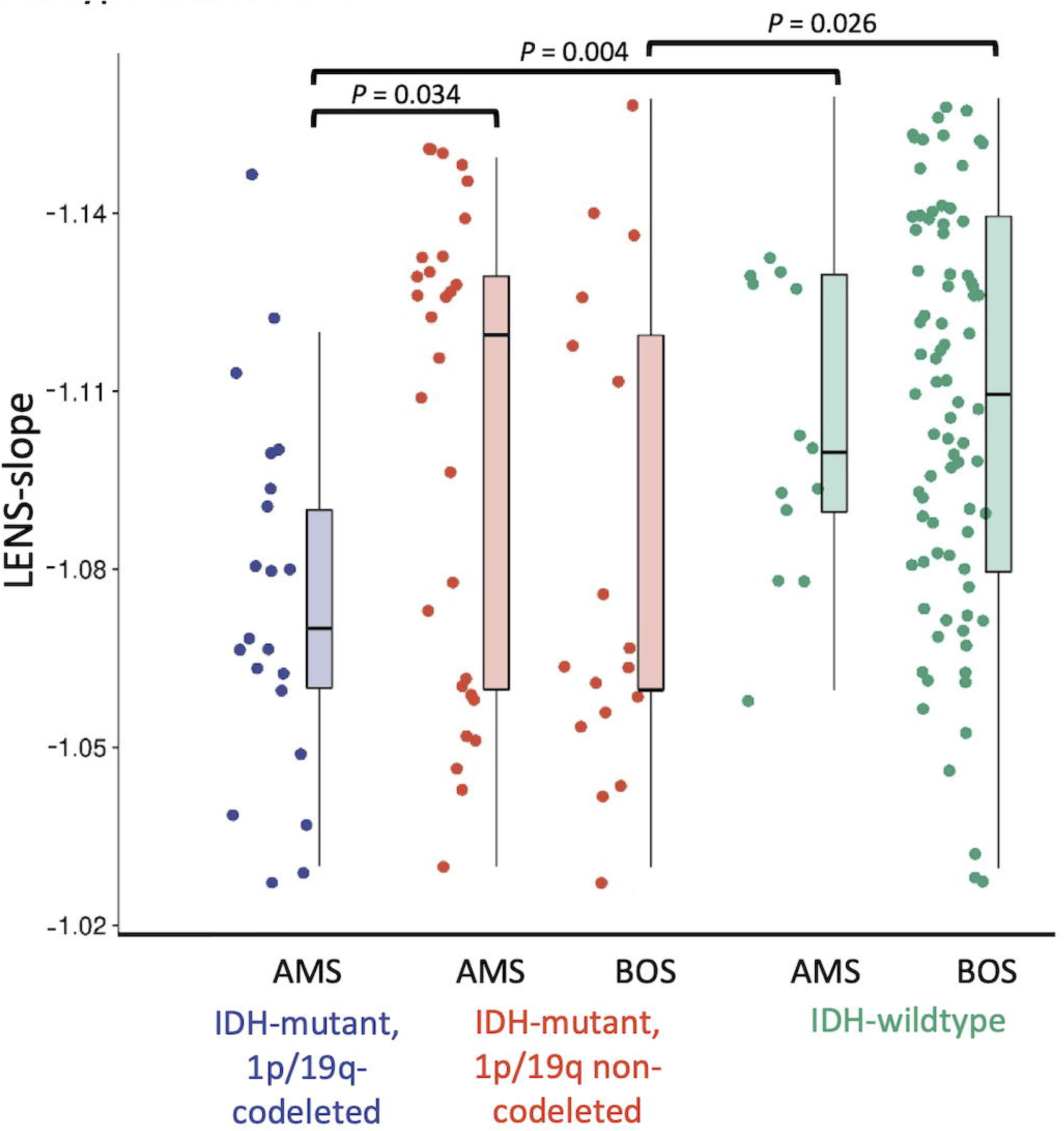
Individual LENS-slope values according to tumor subtype and cohort Legend. This figure shows boxplots and individual values per patient of LENS-slope values for the IDH-mutant, 1p/19q-codeleted gliomas per cohort (left panel). The middle and right panels show the same for the IDH-mutant, 1p/19q non-codeleted gliomas and IDH-wiltype gliomas, respectively. AMS = Amsterdam cohort, MGH = Boston cohort.

When focusing on patients’ performance status, LENS-slope was less steep in patients with a KPS < 90, compared to patients with KPS of 90 or 100 in the AMS cohort (Supplementary Table 3), while LENS-bbp and LENS-offset did not differ between groups. Post-hoc tests revealed that LENS-slope was less steep in patients with KPS < 90 within each subgroup, albeit with significance 0.05 < *P* < 0.1 (Figure 4, Supplementary Table 3). In the MGH cohort, there were no significant differences in LENS between patients with low versus high KPS.

**Figure 4.**
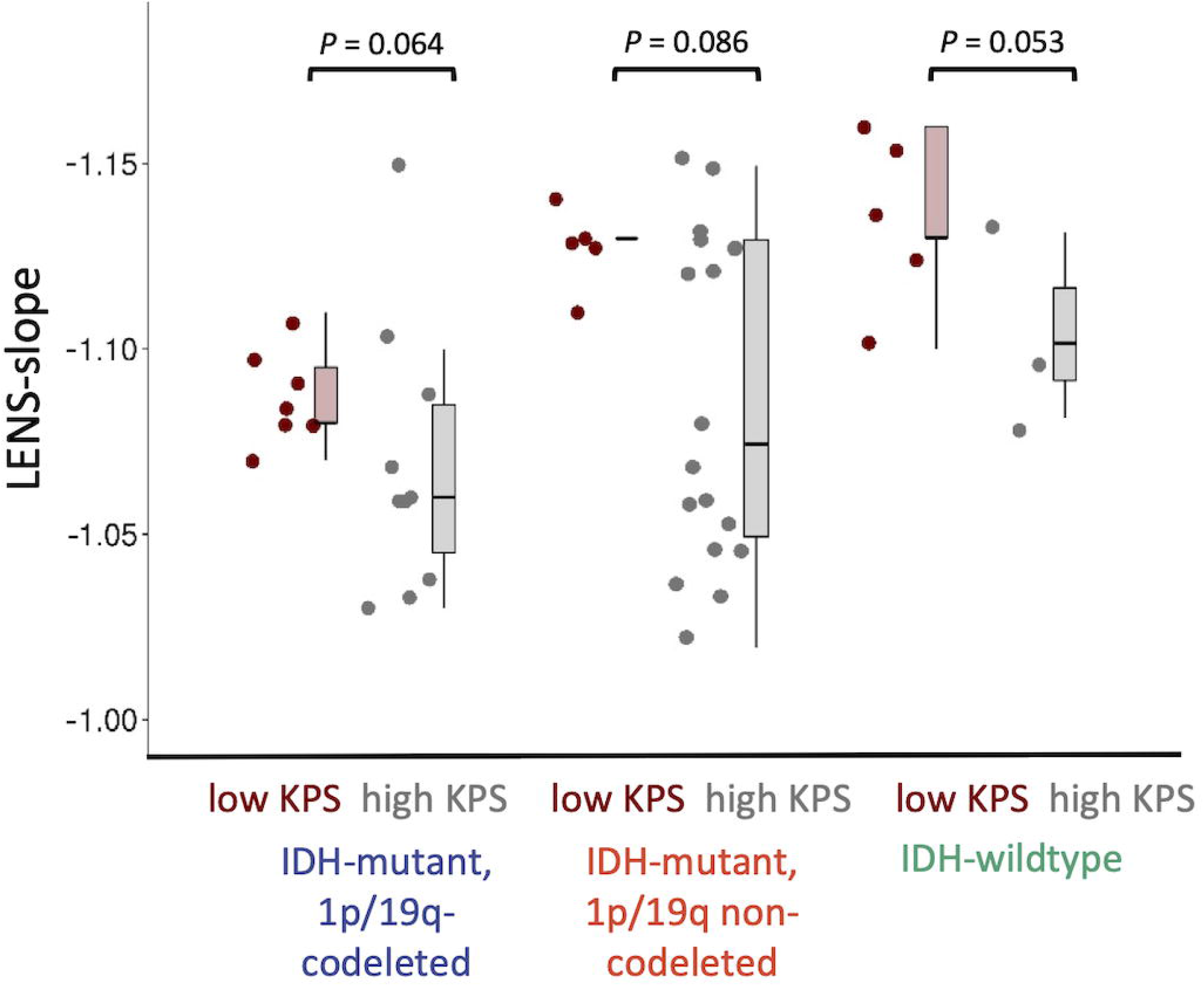
Individual LENS-slope values according to tumor subtype and performance status in the AMS cohort Legend. This figure shows boxplots and individual values per patient of LENS-slope values for the IDH-mutant, 1p/19q-codeleted gliomas in the AMS (Amsterdam) cohort, separated for low (< 90) and high Karnofsky Performance Status (KPS, 90-100) in the left panel. The same is visualized for the IDH-mutant, 1p/19q non-codeleted gliomas (middle panel) and the IDH-wildtype gliomas (right panel).

### Associations between LENS and other clinical variables

All three LENS values were positively related to tumor volume in the IDH-mutant, non-codeleted glioma subgroup of the AMS cohort, also after adjusting for tumor lateralization (Supplementary Table 4). However, these associations were not observed in the smaller MGH cohort, nor were LENS values related to tumor volume in the IDH-mutant, 1p/19q-codeleted and IDH-wildtype gliomas in the AMS cohort.

In the AMS cohort, there were no differences in LENS values between patients with and without epilepsy (Supplementary Table 5). In the MGH cohort, patients with epilepsy had significantly lower LENS-bbp than patients without epilepsy, which proved to be due to the IDH-mutant, non-codeleted subgroup, but not to the IDH-wildtype patients, in post-hoc tests.

The LENS values were not related to PFS or OS in any of the univariate analyses per subgroup (Supplementary Table 6).

## Discussion

We have shown that IDH-mutant, 1p/19q non-codeleted glioma and IDH-wildtype glioblastoma preferentially occur in regions intrinsically characterized by higher brain activity. The most malignant subtype, the IDH-wildtype glioblastoma, was also preferentially localized at regions with a relatively lower intrinsic E/I balance. In contrast, occurrence of the prognostically most favorably subtype, IDH-mutant, 1p/19q-codeleted glioma, did not show any associations with intrinsic regional brain measures. Furthermore, intrinsic E/I balance at each patient’s individual tumor location was lowest in IDH-wildtype glioma patients, followed by IDH-mutant, non-codeleted glioma patients and lastly IDH-mutant, 1p/19q-codeleted glioma patients. Intrinsic E/I balance at individual tumor locations seemed weakly associated with KPS within tumor subgroups, although only in one cohort.

Our hypothesis regarding the preferential occurrence of particularly more malignant subtypes of glioma in brain regions intrinsically characterized by higher neuronal activity was confirmed: higher offset, potentially the best proxy of neuronal spiking rates due to its disentanglement from the oscillatory behavior that is also measured with MEG and captured by broadband power,^41^ was associated with higher occurrence of IDH-mutant, 1p/19q non-codeleted and IDH-wildtype glioma. Preclinical literature describes a causal relationship between peritumoral neuronal spiking rates and tumor growth.^1–6^ Taken together with the current results, regions with intrinsically higher activity may be particularly susceptible to glioma growth, at least in terms of the tumor bulk that is visible on MRI. It remains unclear whether intrinsic spiking rate is a cause of these glioma subtypes originating at particular locations, or merely a contributing factor to its ongoing growth. Regardless of whether brain activity is a cause for occurrence or a byproduct of some underlying mechanism, our results further add to the existing evidence that brain pathology, including glioma, preferentially occurs in brain regions that are intrinsically more active and connected.^14–17,46^ Such hub regions are thought to be more costly and vulnerable due to their metabolic energy demands.^19^ In addition to the recent cellular insights into the complex interplay between the brain and glioma,^10^ these findings underline the circuit-level and potentially whole-brain involvement of healthy brain activity in regional glioma occurrence.

IDH-wildtype glioblastomas also occurred more often in brain regions with intrinsically steeper slope, indicating that a low regional healthy E/I balance (relatively greater inhibition) associates with higher tumor occurrence. In light of the positive correlation between neuronal activity and tumor growth, regions with relatively greater excitation may intuitively seem more likely to harbor these malignant tumors. Previous studies have indicated that alpha band-specific slope is negatively related to offset and power indices when also calculated on narrow frequency bands.^41,42^ However, it is unclear how slope as calculated on the wide range of frequencies as performed here relates to the other measures; our results indicate that greater neuronal activity as measured with power and offset is reflected by relatively higher inhibition over excitation (lower E/I balance). Inhibition is thought to shape cortical activity, potentially to a larger extent than excitation,^47^ particularly when it concerns slower, more distant (>4mm) interactions.^48^ As such, regions with greater potential for inhibition may orchestrate neuronal activity to a larger extent than regions with a higher E/I balance, but future studies combining these measures, investigating both broadband and band-specific components, and verifying the actual contribution of excitatory and inhibitory processes to the signals measured with MEG are necessary.

The regional occurrence of IDH-mutant, 1p/19q-codeleted gliomas did not relate to intrinsic brain activity. This lack of association may be expected: previous preclinical work on the association between neuronal activity and glioma growth has primarily focused on high-grade glioma or glioblastoma,^2,7^ and has also shown that neurogliomal synapses are most likely not present in these codeleted gliomas.^1,49^ Although biologically not yet fully understood, this lack of entanglement between healthy neurons and glioma cells as compared to the non-codeleted gliomas may be reflected in the overall better prognosis that patients with this glioma subtype have. Our findings further support a lack of entanglement and suggest that other mechanisms of tumor growth are at play in these patients, although the small sample size of particularly this subgroup may have obscured associations.

We also considered intrinsic brain activity profiles at individual patients’ tumor locations using LENS, in an effort to translate intrinsic regional activity patterns to clinically relevant tumor and patient characteristics. Particularly LENS-slope robustly differed between tumor subgroups: the IDH-wildtype glioblastoma patients’ values were highest, while IDH-mutant, 1p/19q-codeleted glioma patients’ values were lowest, suggesting that lower E/I balance at individual tumor locations goes hand in hand with increasing malignancy. As such, tumors occurring in regions that are normally characterized by relatively greater inhibitory activity have poorer prognoses. LENS-slope also showed clinical potential, as lower intrinsic E/I balance at the tumor location was found at trend-level in patients with lower KPS, regardless of molecular tumor subgroup. Speculatively, if regions intrinsically favoring greater inhibition orchestrate overall activity and potentially connectivity, are invaded by glioma, integration across different brain regions may be impacted to a larger extent than when the glioma occurs at a location with less inhibitory, integrative activity. We previously reported higher relative inhibitory activity (lower E/I balance) in glioma patients in the regions around the tumor, both as compared to healthy controls and as compared to the contralateral hemishere.^8^ The E/I balance at individual tumor locations may therefore be a relevant factor to further investigate from a clinical perspective, particularly since perampanel, an AMPA inhibitor, has been shown to inhibit tumor growth *in vitro*.^1^ Future work may investigate how intrinsic E/I values at the tumor location relate to pathological decreases in this balance measured at diagnosis, together potentially impacting KPS and perhaps more fine-grained measures of performance, such as cognitive functioning. This is all the more relevant since preserved and dysfunctional activity patterns measured intraoperatively around the tumor during task performance have been shown to be relevant for cognitive processes.^9^

Previous studies have speculated on the role of epilepsy in promoting activity-dependent tumor growth,^4,50^ and seizures were found to directly increase the neurogliomal calcium transients leading to tumor growth and infiltration.^1^ However, we did not find reproducible differences in LENS values between patients with and without epilepsy, potentially due to the low number of patients without epilepsy in our study and the fact that most patients with epilepsy took AEDs. Also, larger tumor volumes did not consistently relate to LENS indices. Of course, tumor volume at diagnosis depends on multiple factors that we cannot disentangle in the current study. For instance, IDH-wildtype gliomas are typically associated with more edema and mass effects due to faster tumor growth, which may lead to faster diagnosis, even at smaller tumor volumes. Longitudinal studies assessing the relationship between tumor volume, LENS values and patients’ own brain activity levels could further elucidate how non-invasively measured brain activity may provide information about tumor volume over time.

Finally, we did not find LENS values to have predictive power in terms of PFS or OS. Previous work has shown that MEG broadband power measured in glioma patients before and after tumor resection relates to prognosis, such that greater broadband power is related to shorter survival.^33,35^ Taking these studies and our current results together, we may speculate that brain activity at the tumor location is already pathologically increased in patients at diagnosis,^33,35^ potentially due to the bidirectional relationship between (intrinsic) neuronal activity and tumor growth. Intrinsic brain activity at the tumor location, as measured in healthy controls like in the current work, may therefore hold no specific prognostic value, but longitudinal studies should further elucidate how brain activity changes during the course of the disease. Faster disease trajectories could for instance be expected for tumors located in brain areas with intrinsically higher brain activity and that also have larger pathological increases of activity at diagnosis.

Several limitations should be kept in mind when interpreting these results. First of all, sample sizes of subgroups, particularly the IDH-mutant, 1p/19q-codeleted patients, were relatively small and may have obscured results. Also, particularly the AMS cohort may have suffered from selection bias, as these patients underwent task-based fMRI and may not represent the complete population of glioma patients. Secondly, direct translation of preclinical work to our patient study is hampered by several factors. We non-invasively measured neuronal activity, which may have impacted the comparability between previous work concerning spiking rates and the current results. Thirdly, particularly the slope of the aperiodic component of the power spectrum, thought to reflect the balance between excitation and inhibition of the underlying neuronal populations, is a novel measure and has not been extensively validated. Moreover, we adhered to several choices made in previous work regarding the frequency range of interest and parameters used for the analysis of the offset and slope, for which currently no gold standard exists. Finally, we used intrinsic brain activity indices by averaging regional indices over a cohort of healthy controls, although it is clear that there is variation across these subjects that we did not take into account in our analyses. Furthermore, our calculation of LENS values was based on the weighted overlap between patients’ individual tumor masks and average regional brain activity values, while the contribution of different atlas regions’ intrinsic activity values is likely to be more complex.

In conclusion, we have shown that more malignant glioma occurs more often in brain regions that are intrinsically more active, with the most malignant glioblastomas also favoring areas that are intrinsically marked by a lower E/I balance. We also described how intrinsic E/I balance at patients’ individual tumor locations independently relates to tumor subtype and performance status, such that patients with prognostically unfavorable tumor subtypes and poorer KPS have tumors in regions with relatively higher inhibition. Taken together, our findings offer a better understanding of activity-dependent preferential tumor occurrence and indicate that glioma localization in terms of intrinsic brain activity levels reflects both tumor biology and performance status.

## Data Availability

The derivative data and code used in this manuscript are available.

https://github.com/multinetlab-amsterdam/projects/

## Supplementary material

There are Supplementary Materials associated with this manuscript.

## Abbreviations

Bbp: broadband power
LENS: gLioma-spEcific braiN activity Signature
MEG: Magnetoencephalography

