## Supplementary for "Regional healthy brain activity, glioma occurrence and symptomatology"

### Supplementary Materials

#### Spin test results

Supplementary Table 1. Associations between regional glioma occurence and regional intrinsic brain activity using the spin test

| Subgroup | Cohort | N | Broadband power | Offset | Slope |
|---|---|---|---|---|---|
| IDH-mutant, non-codeleted | AMS | 30 | NA | 0.396 (0.023)* | NA |
| | MGH | 18 | NA | 0.305 (0.137) | NA |
| | *combined* | *48* | *NA* | *0.407 (0.029)** | *NA* |
| IDH-wildtype glioma | AMS | 14 | NA | 0.418 (0.051) | NA |
| | MGH | 91 | 0.278 (0.006)* | 0.315 (0.034)* | -0.437 (0.005)* |
| | TCGA | 102 | 0.270 (0.003)* | 0.309 (0.040)* | -0.387 (0.002)* |
| | *combined* | *207* | *0.307 (0.004)** | *0.429 (0.015)** | -0.446 (0.010)* |

Cells indicate Spearman correlation (P-value). * $P < 0.05$ (note that correlation coefficients are the original correlations as indicated in Table 2. AMS = Amsterdam, MGH = Boston, TCGA = The Cancer Genome Atlas. Values indicate Spearman correlation coefficients.

#### Impact of medial regions

As evident from Supplementary Figure 1 on the next page, medial regions tended to have higher brain activity values in healthy controls. Since areas further away from the MEG sensors are known to have lower signal-to-noise (SNR) ratios than regions on the cortical surface,[1] we cannot fully exclude the potential confounding effect of SNR on brain activity values from those regions. Furthermore, since glioma also often occurs more medially, in the white matter, than on the cortical surface, where the MEG signal originates,[2] we used only the regions with brain activity values below the top quartile (i.e. regions with values lower than the 75$^{th}$ percentile) for our main analysis. When taking all regions into account, however, all significant main results are replicated (Supplementary Table 2).

Supplementary table 2. Associations between regional glioma occurrence and regional intrinsic brain activity when taking all regions into account

| **Subgroup** | **Cohort** | **N** | **Broadband power** | **Offset** | **Slope** |
|---|---|---|---|---|---|
| IDH-mutant, 1p/19q-codeleted glioma | *AMS* | *21* | *-0.059* | *-0.010* | *0.239** |
| IDH-mutant, non-codeleted glioma | AMS | 30 | 0.188 | 0.433* | -0.258* |
| | MGH | 18 | 0.051 | 0.267* | -0.002 |
| | *combined* | *48* | *0.177* | *0.424** | *-0.208** |
| IDH-wildtype glioma | AMS | 14 | 0.057 | 0.282* | -0.211 |
| | MGH | 91 | 0.272* | 0.335* | -0.373* |
| | TCGA | 102 | 0.243* | 0.287* | -0.440* |
| | *combined* | *207* | *0.233** | *0.358** | *-0.460** |

* $P < 0.0019$, significant after correction for multiple comparisons across 27 tests. AMS = Amsterdam, MGH = Boston, TCGA = The Cancer Genome Atlas. Values indicate Spearman correlation coefficients.

Supplementary Figure 1. Average regional brain activity and variability in healthy controls

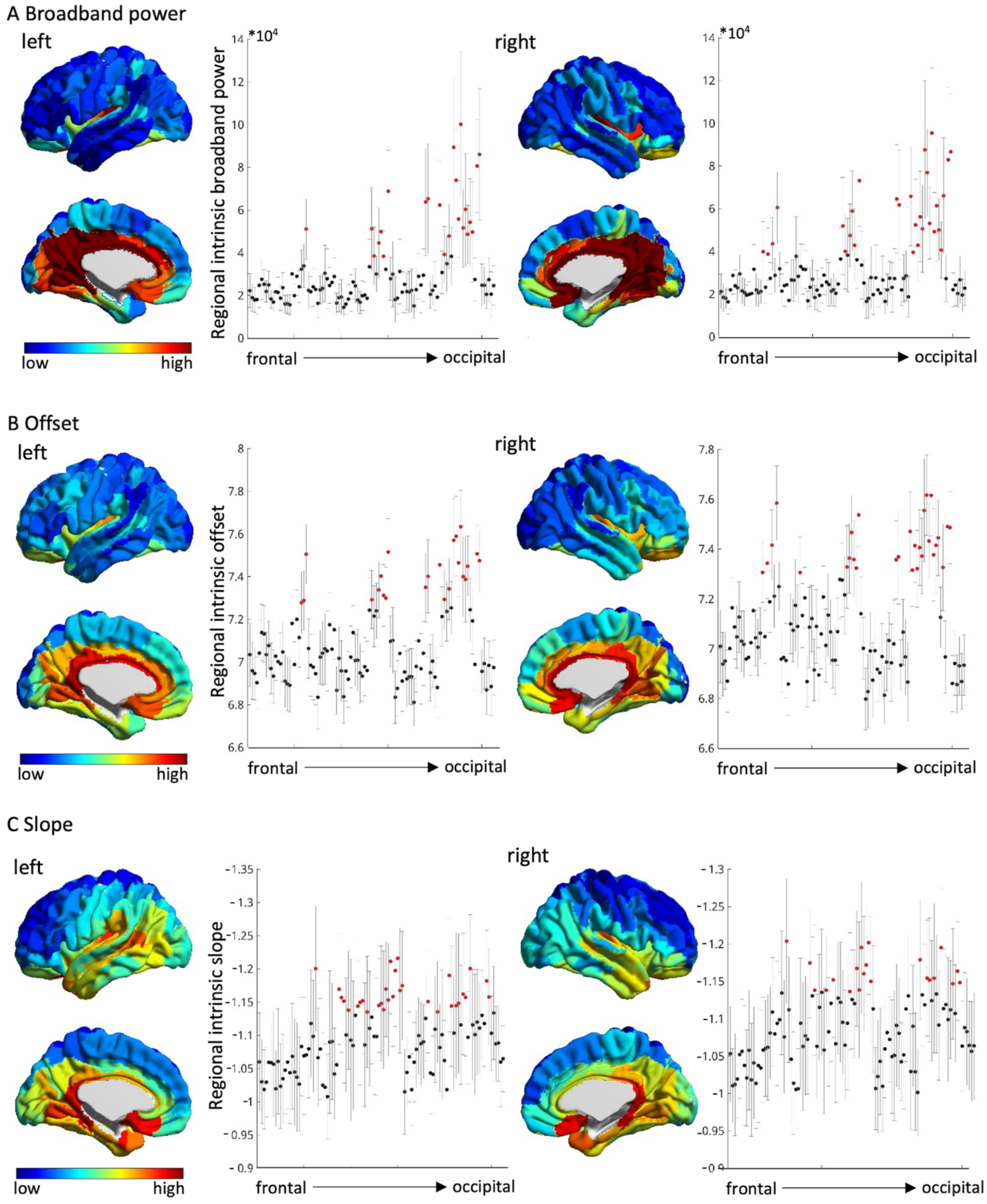


Legend. (A) depicts the spatial distribution of average broadband power plotted on the brain for the left and right lateral and medial hemispheres, in addition to the variability of regional broadband power values, indicated by gray error bars that represent standard deviation over the 45 healthy controls included in the analysis. Black dots are the non-medial regions, red dots are the medial regions that are subject to lower signal-to-noise ratio and were excluded from our primary analyses. (B) displays the same for offset, and (C) for slope.

Supplementary Figure 2. Tumor occurrence maps per cohort

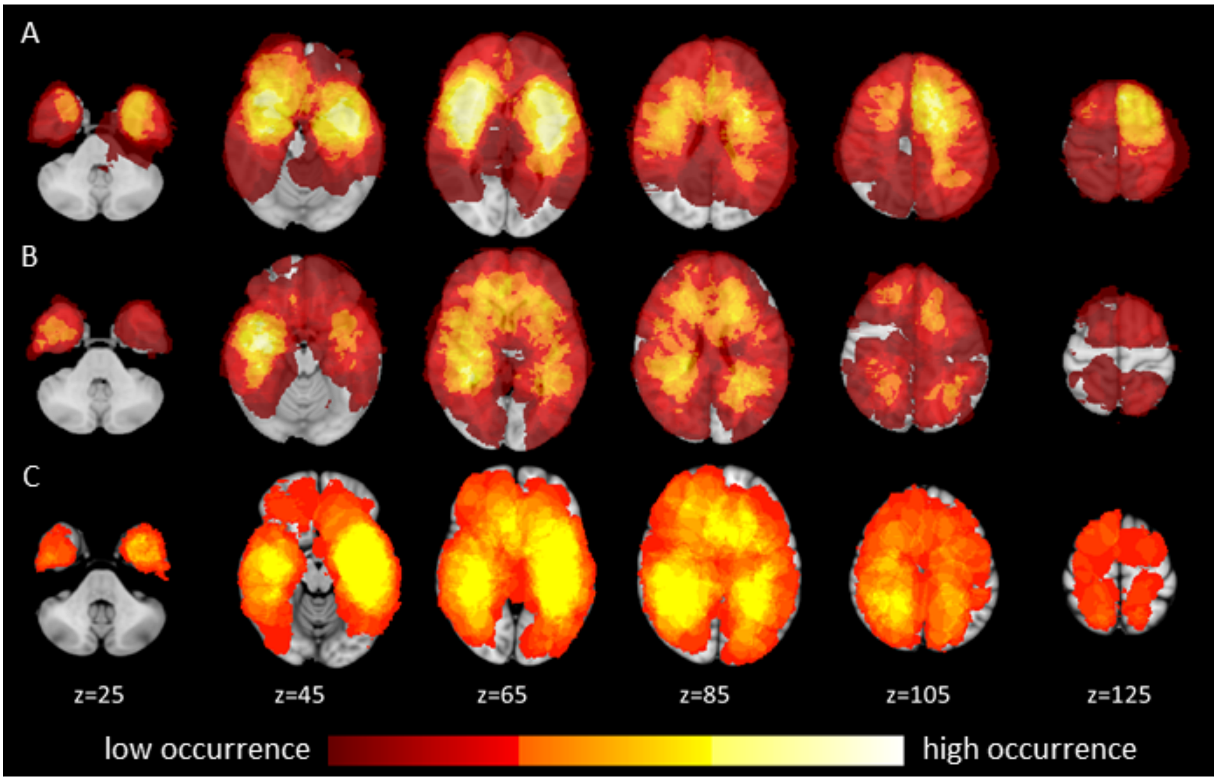


Legend. Tumor occurrence maps for (A) the Amsterdam (AMS) cohort, (B) the Massachusetts General Hospital (MGH) cohort, and (C) The Cancer Genome Atlas (TCGA) cohort.

### Impact of age of healthy controls

The included healthy controls were matched to the primary AMS cohort and were significantly younger than patients in the MGH cohort ($t(152) = 5.035$, $P < 0.001$). Because of this age difference, additional analyses were performed to assure that any findings specifically in the MGH/TCGA cohorts and the IDH-wildtype gliomas were not due to this age difference. We calculated average regional brain activity values for the youngest healthy controls (age < 45 years, n = 21) and the oldest healthy controls (age > 45 years, n = 44) separately, to explore the potential confounding effects of this age difference on our analyses.

Results show that the distribution of regional brain activity was nearly identical between these two healthy control groups (broadband power Spearman's rho = 0.973, $P < 0.001$; offset Spearman's rho = 0.983, $P < 0.001$; Spearman's slope rho = 0.965, $P < 0.001$). We therefore decided to generally use the same 45 healthy controls for all analyses, to avoid comparisons with different control groups for each subgroup. However, for specific analyses within the older IDH-wildtype glioblastoma patients, we replicated our main findings using only the better matched, older healthy controls (data not shown).

### Regional versus individual variability

Since we used the average of regional intrinsic activity values over all controls as either a correlate of tumor occurrence or to construct signatures of activity according to individual patients' tumor locations, we further explored the spatial distribution and variation around the mean of the three brain activity values (see Supplementary Figure 1). These figures suggest that there are meaningful differences between brain regions with respect to intrinsic activity values. Particularly for offset and slope, there are variations in the average values along the anterior-posterior axis that exceed the variability over controls, also in those non-medial regions we included in our main analyses. For broadband power, there are less consistent patterns to be seen, although the effect of lower SNR in the medial regions (also indicated by the greater variability for those red dots) may have obscured some variations in the non-medial areas.

Supplementary table 3. Differences in brain activity between patients with low and high KPS

| | AMS cohort | | | |
|---|---|---|---|---|
| **Brain activity measure** | **KPS < 90 (n=23)** | **KPS 90 or 100 (n=38)** | **test statistic** | **p-value** |
| LENS-bbp | 33034 (28378-37306) | 31392 (24616-38882) | U = 371 | 0.326 |
| LENS-offset | 7.17 (7.06-7.25) | 7.15 (7.02-7.26) | U = 387 | 0.457 |
| LENS-slope | 1.11 (1.08-1.13) | 1.08 (1.05-1.13) | U = 299 | 0.040* |
| **Subgroup (LENS-slope only)** | **KPS < 90** | **KPS 90 or 100** | **test statistic** | **p-value** |
| IDH-mutant, 1p/19q-codeleted | n=7; 1.08 (1.08-1.11) | n=10; 1.06 (1.04-1.09) | U = 16 | 0.064 |
| IDH-mutant, non-codeleted | n=5; 1.13 (1.12-1.14) | n=18; 1.07 (1.05-1.13) | U = 22 | 0.086 |
| IDH-wildtype | n=5; 1.13 (1.12-1.16) | n=3; 1.10 (1.08-1.10) | U = 1 | 0.053 |

| | MGH cohort | | | |
|---|---|---|---|---|
| **Brain activity measure** | **KPS < 90 (n=29)** | **KPS 90 or 100 (n=66)** | **test statistic** | **p-value** |
| LENS-bbp | 31983 (27344-36872) | 30907 (25320-40533) | U = 884 | 0.555 |
| LENS-offset | 7.15 (7.06-7.22) | 7.13 (7.02-7.23) | U = 843 | 0.357 |
| LENS-slope | 1.12 (1.08-1.14) | 1.10 (1.07-1.14) | U = 852 | 0.396 |

* $P < 0.05$. Cells indicate median (Q1-Q3), and subgroup sample size where necessary. KPS = Karnofsky Performance Scale

Supplementary Table 4. Associations between LENS and tumor volume per subgroup and cohort, adjusted for tumor lateralization

| **(Sub)group** | **Cohort** | **n** | **Broadband power** | **Offset** | **Slope** |
|---|---|---|---|---|---|
| Subgroup 1 (IDH-mutant, 1p/19q-codeleted) | AMS | 21 | 0.256 | 0.299 | -0.339 |
| Subgroup 2 (IDH-mutant, non-codeleted) | AMS | 30 | 0.371* | 0.458* | -0.478** |
| | MGH | 18 | -0.275 | -0.149 | 0.033 |
| Subgroup 3 (IDH-wildtype) | AMS | 14 | -0.088 | 0.010 | 0.285 |
| | MGH | 91 | 0.078 | 0.117 | 0.047 |

* $P < 0.05$, ** $P < 0.01$. AMS = Amsterdam, MGH = Boston. Values indicate standardized betas.

Supplementary table 5. Differences in LENS values between patients with and without epilepsy

| | | AMS cohort | | |
|---|---|---|---|---|
| **Brain measure** | **Epilepsy (n=60)** | **No epilepsy (n=11)** | **test statistic** | **p-value** |
| LENS-bbp | 31675 (25750-37783) | 29692 (22262-35330) | U = 388 | 0.357 |
| LENS-offset | 7.16 (7.02-7.25) | 7.13 (7.03-7.15) | U = 391 | 0.332 |
| LENS-slope | -1.11 (-1.06--1.13) | -1.08 (1.06--1.13) | U = 402 | 0.253 |

| | | MGH cohort | | |
|---|---|---|---|---|
| **Brain measure** | **Epilepsy (n=69)** | **No epilepsy (n=22)** | **test statistic** | **P-value** |
| LENS-bbp | 31341 (25388-38801) | 56335 (26367-64838) | U = 544 | 0.046* |
| LENS-offset | 7.12 (7.03-7.22) | 7.18 (7.04-7.39) | U = 563 | 0.069 |
| LENS-slope | -1.12 (-1.07--1.14) | -1.12 (-1.08--1.15) | U = 690 | 0.522 |
| **LENS-bbp only** | **Epilepsy** | **No epilepsy** | **test statistic** | **P-value** |
| IDH-mutant, non-codeleted | n=12; 26368 (23037-40426) | n=5; 37030 (29940-75994) | U = 9 | 0.027* |
| IDH-wildtype | n=57; 32506 (25690-41788) | n=17; 35640 (26079-59489) | U = 398 | 0.266 |

* p < 0.05. Cells indicate median (Q1-Q3), and subgroup sample size where appropriate.

Supplementary table 6. Associations between LENS values and survival

| | | AMS cohort | | |
|---|---|---|---|---|
| **IDH-mutant, 1p/19q-codeleted** | **PFS** | **p-value** | **OS** | **p-value** |
| LENS-bbp | 1.00 (1.00-1.00) | 0.947 | 1.00 (1.00-1.00) | 0.639 |
| LENS-offset | 0.31 (0.01-74.58) | 0.676 | 11.26 (0.01-2.77*$10^5$) | 0.639 |
| LENS-slope | 0.47 (0.01-2.10*$10^7$) | 0.933 | 4.55 (0.01-2.16*$10^{15}$) | 0.930 |
| **IDH-mutant, non-codeleted** | **PFS** | **p-value** | **OS** | **p-value** |
| LENS-bbp | 1.00 (1.00-1.00) | 0.742 | 1.00 (1.00-1.00) | 0.879 |
| LENS-offset | 1.45 (0.06-35.56) | 0.821 | 1.94 (0.01-276.10) | 0.793 |
| LENS-slope | 0.319 (0.01-92.43*$10^2$) | 0.827 | 95.08 (0.01-6.20*$10^9$) | 0.620 |
| **IDH-wildtype** | **PFS** | **p-value** | **OS** | **p-value** |
| LENS-bbp | 1.00 (1.00-1.00) | 0.430 | 1.00 (1.00-1.00) | 0.363 |
| LENS-offset | 5.81 (0.07-478.16) | 0.434 | 529 (0.04-642.65) | 0.496 |
| LENS-slope | 1373.42 (0.01-9.30*$10^{10}$) | 0.432 | 8.40 (0.01-2.01*$10^9$) | 0.829 |
| | | **MGH cohort** | | |
| **IDH-mutant, non-codeleted** | **PFS** | **p-value** | **NA** | **NA** |
| LENS-bbp | 1.00 (1.00-1.00) | 0.467 | | |
| LENS-offset | 16.28 (0.02-16453.74) | 0.429 | | |
| LENS-slope | 245.63 (0.01-1.24*$10^{11}$) | 0.590 | | |
| **IDH-wildtype** | **PFS** | **p-value** | **OS** | **p-value** |
| LENS-bbp | 1.00 (1.00-1.00) | 0.071 | 1.00 (1.00-1.00) | 0.861 |
| LENS-offset | 3.35 (0.79-14.15) | 0.100 | 0.91 (0.19-4.26) | 0.902 |
| LENS-slope | 28.84 (0.05-16.47*$10^3$) | 0.299 | 1.67 (0.01-830.06) | 0.871 |

Cells indicate HR (95% confidence interval). PFS = progression-free survival, OS = overall survival. Note that none of the MGH IDH-mutant, non-codeleted patients died within follow-up.
